# The third study of infectious intestinal disease in the community microbiological methods

**DOI:** 10.1101/2025.11.20.25340668

**Authors:** C. Bronowski, A. Hughes, C. Jenkins, G. Godbole, R. Chalmers, C. Celma, S. Beard, M. McDowell, M. Oates, K. Elwin, E. Cunningham-Oakes, A.C Darby, D. Hungerford, A. Mill, S. O’Brien, M. Hopkins, N.A. Cunliffe, the IID3 Consortium

## Abstract

**Introduction:** Infectious intestinal disease (IID) is a significant public health concern, with an estimated 17 million cases occurring annually in the UK. However, only a small proportion of cases in the community present to primary health care, and those that do may not be tested for the causative pathogen. The third study of IID in the community (IID3 study) will provide updated estimates of the incidence and aetiology of IID in the UK. This protocol describes the laboratory methods employed by the IID3 study in primary diagnostic and reference laboratories.

**Methods and analysis:** Stool specimens obtained from symptomatic subjects enrolled in two cohorts recruited prospectively through Primary Care in the UK are tested for 21 pathogens using PCR-based methods, in two separate workflows at Liverpool Clinical Laboratories. The first workflow comprises EntericBio DX, Viral and *Clostridium difficile* assays (Serosep, Crawley, UK); and the second a panel of assays for detection of *Clostridium perfringens, Aeromonas spp., Cyclospora cayetanensis* and diarrhoeagenic *Escherichia coli* based on 3base® Technology (Genetic Signatures, Newtown, Australia). Specimens testing positive for bacterial pathogens are subjected to culture. Specimens positive for, and isolates of *Salmonella, Campylobacter, Yersinia, Vibrio* and *Shigella* species and diarrhoeagenic *E. coli* are referred to the Gastrointestinal Bacteria Reference Unit, UK Health Security Agency (UKHSA) for whole genome sequencing and antimicrobial susceptibility testing. The parasites *Cryptosporidium*, *Giardia* and *Cyclospora* are referred for genotyping to the national Cryptosporidium Reference Unit, Public Health Wales. Norovirus positive faecal specimens are referred for Sanger Sequencing-based genotyping to the Enteric Virus Unit, UKHSA.

**Ethics and dissemination:** Favourable ethical opinion was granted on the 10^th^ of August 2022 by the East Midlands - Nottingham 1 Research Ethics Committee (IRAS ID 314268, REC Reference 22/EM/0130). The results will be disseminated to key stakeholders including the Food Standards Agency, participating general practices and to the academic community through publications and conference presentations. A real-time breakdown of pathogens identified in faecal samples could be viewed openly on the study [Dashboard](https://app.powerbi.com/view?r=eyJrIjoiNjZiODEwMmUtNWU4Yi00YzJlLTliNjgtYmE1NzA5YWNmNDhmIiwidCI6ImNjOTVkZTFiLTk3ZjUtNGY5My1iNGJhLWZlNjhiODUyY2Y5MSJ9) throughout the surveillance period.

**Article Summary:** *Strengths and limitations of this study:*

- A total of 5439 stool samples underwent molecular testing for a total of 21 organisms at the Liverpool Clinical Laboratories (LCL), with detailed pathogen characterisation at national reference laboratories.
- A commercial Genetic Signatures assay was evaluated in a routine diagnostic setting using commercially-provided pathogen controls, reference isolates and clinical samples, enabling the inclusion of *Aeromonas* and ETEC PCR targets into the diagnostic panel.
- Real-time referrals of PCR-reactive samples were made to UK Health Security Agency (UKHSA) regardless of bacterial culture results, and to Public Health Wales (PHW).
- Norovirus PCR-reactive samples were referred to the Enteric Virus Unit at UKHSA for genotyping ahead of Norovirus becoming a notifiable infection on the 6^th^ of April 2025, adding valuable information on Norovirus genotypes circulating in the general population.
- A Biobank has been created for residual faecal specimens, forming a rich resource for future studies.
- A major challenge was the lack of a harmonised UK-wide digital referral and reporting system, resulting in an inflated manual reporting workload for staff at LCL.

## Introduction

Infectious Intestinal Disease (IID) is a common illness in the UK population, with an estimated 17 million cases reported per year [1]. Although symptoms of IID are generally mild and self-limiting, clinical outcomes can be severe, including bloody diarrhoea, abdominal pain, fever, nausea and vomiting, and in the most severe cases IID can result in hospitalisation and death. The economic and clinical burden of IID in England is high, with 20-25% of the population experiencing symptoms each year, among whom half report taking days off work or school [1].

IID is caused by a wide range of causative organisms, including bacteria (e.g. *Salmonella spp.* and *Campylobacter spp.*), viruses (e.g. Norovirus and Rotavirus) and parasites (e.g. *Giardia duodenalis* and *Cryptosporidium spp.*). Pathogens causing IID can be spread through contaminated food, water, fomites, or by person-to-person or animal-to-person contact. The purpose of microbiological surveillance is to identify the microorganisms causing IID, and their source and transmission pathways, with the aim of controlling, reducing and ultimately preventing infection through targeted interventions. Surveillance of IID in the United Kingdom falls under the remit of the UK Health Security Agency (UKHSA) in England, Public Health Agency (PHA) in Northern Ireland, Public Health Wales (PHW) and Public Health Scotland (PHS), which monitor clinical notifications, laboratory-based reporting of cases, outbreaks, and hospital data to understand the burden and trends of IID. However, national surveillance relies on laboratory reports of pathogens identified in stool specimens from community and hospital patients reporting symptoms of IID, and this captures only a fraction of the total cases [1].

Two UK-based studies of similar design conducted between 1993-1996 (IID1) [2] and then between 2007-2009 (IID2) [1] highlighted significant under-reporting of IID cases, which were often not assigned to a causative organism. The IID1 study showed a large under-ascertainment of IID in national surveillance, since for every one case reported to national surveillance six cases presented to GPs and 23 cases were present in the community; the most commonly identified bacterial pathogen was *Campylobacter*, while the most commonly identified viruses were Norovirus and Rotavirus (parasites were not sought) [2]. IID1 highlighted a need for improved surveillance and better diagnostics, thereby influencing public health policy and investment in laboratory capacity. The IID2 study saw the advent of molecular diagnostic testing with the introduction of PCR and inclusion of parasites, demonstrating that the burden of IID remained high with Norovirus emerging as a leading cause of IID, particularly in adults; *Campylobacter* remained the commonest bacterial pathogen. IID2 supported the expansion of molecular testing in diagnostic laboratories, reaffirmed the continued gap between community cases and national surveillance data and provided a benchmark for monitoring disease burden over time [1].

Diagnostic microbiology has made significant advances since IID2, with molecular techniques such as commercial PCR-based panels and Whole Genome Sequencing (WGS) being routinely used in both diagnostic and reference laboratories, although there is incomplete uptake of PCR in diagnostic laboratories. The availability of WGS offers enhanced information on pathogens such as circulating genotypes, bacterial virulence markers and antimicrobial resistance-gene profiles. Here we describe the diagnostic and reference laboratory testing employed by IID3 to generate updated knowledge on the causes of IID in the UK.

## Methods and analysis

### Study Overview

The IID3 study (2023-2026), comprises three community-based cohorts: a population-based prospective cohort and two prospective cohorts in Primary Care, which are described elsewhere [IID3 protocol paper, *personal communication*]. Primary diagnostic testing of stool specimens is undertaken at LCL, using molecular panels that target an extended spectrum of causative agents, compared to routine diagnostics. Reference microbiological investigation of bacterial isolates and faecal specimens is undertaken at the UKHSA Gastrointestinal Bacteriology Reference Unit (bacteria) and Enteric Virus Unit (viruses) and Public Health Wales, Cryptosporidium Reference Laboratory (parasites).

### Public and Patient Involvement

The study has public and patients’ involvement (PPI) at all stages. We have input from Patient Participation Groups in primary care, the PPI research theme and panel from the NIHR Health Protection Research Unit in Gastrointestinal Infections and PPI representatives on the IID3 Study Executive Committee and the External Advisory Panel. We have incorporated feedback on how to phrase questions about sex/gender, and the reading age of material for children. We have received feedback from a Primary School teacher who thinks that the material should be suitable for a reading age of 7 to 8 years. We have also incorporated feedback from older members of the community to make sure that the material is unlikely to cause offence. PPI involvement will continue throughout the lifecycle of the study.

A public facing dashboard ([IID3dashboard](https://app.powerbi.com/view?r=eyJrIjoiNjZiODEwMmUtNWU4Yi00YzJlLTliNjgtYmE1NzA5YWNmNDhmIiwidCI6ImNjOTVkZTFiLTk3ZjUtNGY5My1iNGJhLWZlNjhiODUyY2Y5MSJ9)) reporting on the pathogens identified and GP practices recruited is updated in real time.

The FSA hosts a designated IID3 website ([FSA_IID3](https://www.food.gov.uk/research/foodborne-pathogens/the-third-study-of-infectious-intestinal-disease-in-the-uk-iid3)) with further information and updates about the study, such as a seminar about the study and Blog posts ([food.blog.gov.IID3](https://food.blog.gov.uk/2024/09/16/how-technology-has-enabled-large-scale-rapid-population-level-recruitment-into-research-possible-food-poisoning-in-the-community-the-third-infectious-intestinal-disease-study-iid3/), [science.food.gov.IID3](https://science.food.gov.uk/post/3525-uncovering-the-hidden-burden-of-infectious-intestinal-disease-early-insights-from-iid3)).

Once the study has been published, participants will be informed of the results through the FSA website and potential news items in other outlets. Data will also flow into the UKHSA reporting.

### Case definition

IID3 employs the same case definition used for the earlier IID studies to ensure comparability. Cases are defined as individuals experiencing loose stools or clinically significant vomiting lasting less than two weeks, with no known non-infectious cause and preceded by a symptom-free period of at least three weeks. Vomiting is deemed clinically significant if it occurred more than once within 24 hours and either caused incapacitation or was accompanied by additional symptoms such as abdominal cramps or fever. Data are collected so as to be able to apply the international case definition developed by Majowicz *et al* [3], where a case of gastroenteritis is defined as an individual with 3 or more loose stools, or any vomiting, in 24 hours which is unrelated to existing health conditions, medication use, or other non-infectious causes.

### Cohorts

This study comprised three cohorts with participant recruitment and record-linked access to primary care routine data:

Cohort 1 (Prospective Cohort Study) is a population-based household cohort, to identify the incidence of IID in the community that may not present to general practice.

Cohort 2 (GP Presentation study) is a prospective cohort of patients presenting to primary care with symptoms of IID.

Stool specimens are collected for IID cases in cohorts 1 and 2. Contemporaneous anonymous results for cohorts 1 and 2 are shown on a publicly available [online dashboard](https://app.powerbi.com/view?r=eyJrIjoiNjZiODEwMmUtNWU4Yi00YzJlLTliNjgtYmE1NzA5YWNmNDhmIiwidCI6ImNjOTVkZTFiLTk3ZjUtNGY5My1iNGJhLWZlNjhiODUyY2Y5MSJ9).

Cohort 3 (GP Enumeration study) provides contemporary data about routine practice through a 12-month prospective audit of IID cases presenting to GPs, observing cases as they presented in normal clinical practice. Stool specimen submission and testing is undertaken as per routine practice in local/regional laboratories, so is not described here.

### Participant enrolment

Participant consent to enrol in the study is obtained for cohorts 1 and 2 through a mobile app, together with collection of demographic and IID symptom data. For cohort 1, general practices used the portal to view when participants reported illness on their weekly symptom questionnaires and to send an automated email asking participants to collect a stool specimen kit [4].

### Stool microbiology for cohorts 1 and 2

Commercial PCR assays supplied by Serosep (Crawley, UK) and Genetic Signatures (Newtown, Australia) are employed for the qualitative detection of pathogen nucleic acid in stool specimens (see box 1 for specific kits used and organisms sought). All specimens are tested within 24 hours of laboratory receipt using Serosep EntericBio DX, Viral and Clostridioides difficile kits for simultaneous detection of 15 bacteria, viruses and parasites of clinical interest. Stool is pre-treated with EntericBio Stool Preparation Solution prior to batch PCR setup using the EntericBio workstation followed by amplification and detection on the Roche LightCycler 480 II (Roche Diagnostics, Burgess Hill, UK). Data analysis is standardised using FastFinder Software (UgenTec). Specimens testing positive for *Campylobacter*, *Salmonella*, *Shigella*, STEC, *Vibrio* or *Yersinia* by PCR are selected for routine culture and identification as detailed in Table 1. The C. diff Quik Chek Complete® rapid immunoassay (Techlab, USA) is used for toxin detection in specimens with reactive EntericBio *C. difficile* results.

**Table 1.** Routine reflex investigations for samples with reactive EntericBio results.

| EntericBio Result | Sub-Culture media | Further Tests |
| --- | --- | --- |
| **Salmonella reactive** | **XLD**<br>O<sub>2</sub> 16-24hr at 35-37°C<br>**SELENITE F**<br>O<sub>2</sub> 16-24hr at 35-37°C then sub XLD if primary XLD is negative. | MALDI, API20E, Serology, Sensitivities and refer to Reference Laboratory. |
| **Shigella reactive** | **XLD**<br>O<sub>2</sub> 16-24hr at 35-37°C | MALDI, API20E, Serology, Sensitivities and refer to Reference Laboratory |
| **Campylobacter reactive** | **CAMP**<br>Micro-aerobic<br>40-48hr at 39-42°C | MALDI and sensitivities, refer to Reference Laboratory |
| **STEC reactive** | **SMAC**<br>O<sub>2</sub> 16-24hr at 35-37°C | MALDI, Serology, Sensitivities and refer to Reference Laboratory. |
| **Yersinia reactive** | **Yersinia selective**<br>O<sub>2</sub> 24-48hr at 28-30°C | MALDI and refer to Reference Laboratory. |
| **Vibrio reactive** | **TCBS & APW**<br>O<sub>2</sub> 16-24hr at 35-37°C<br>Day2: Sub 1 loop-full from APW to 2nd TCBS if primary TCBS is negative. | MALDI, Serology and refer to Reference Laboratory. (Cholerae and Parahaemoliticus.) |
| **Giardia reactive** | Not applicable | Aliquot of original stool specimen referred to Cryptosporidium Reference Unit. |
| **Cryptosporidium reactive** | Not applicable | Aliquot of original stool specimen referred to Cryptosporidium Reference Unit. |
| **E. histolytica reactive** | Not applicable | No further action required. |
| **Clostridioides difficile reactive** |  | Perform Quik Check Toxin test |

**Box 1.**
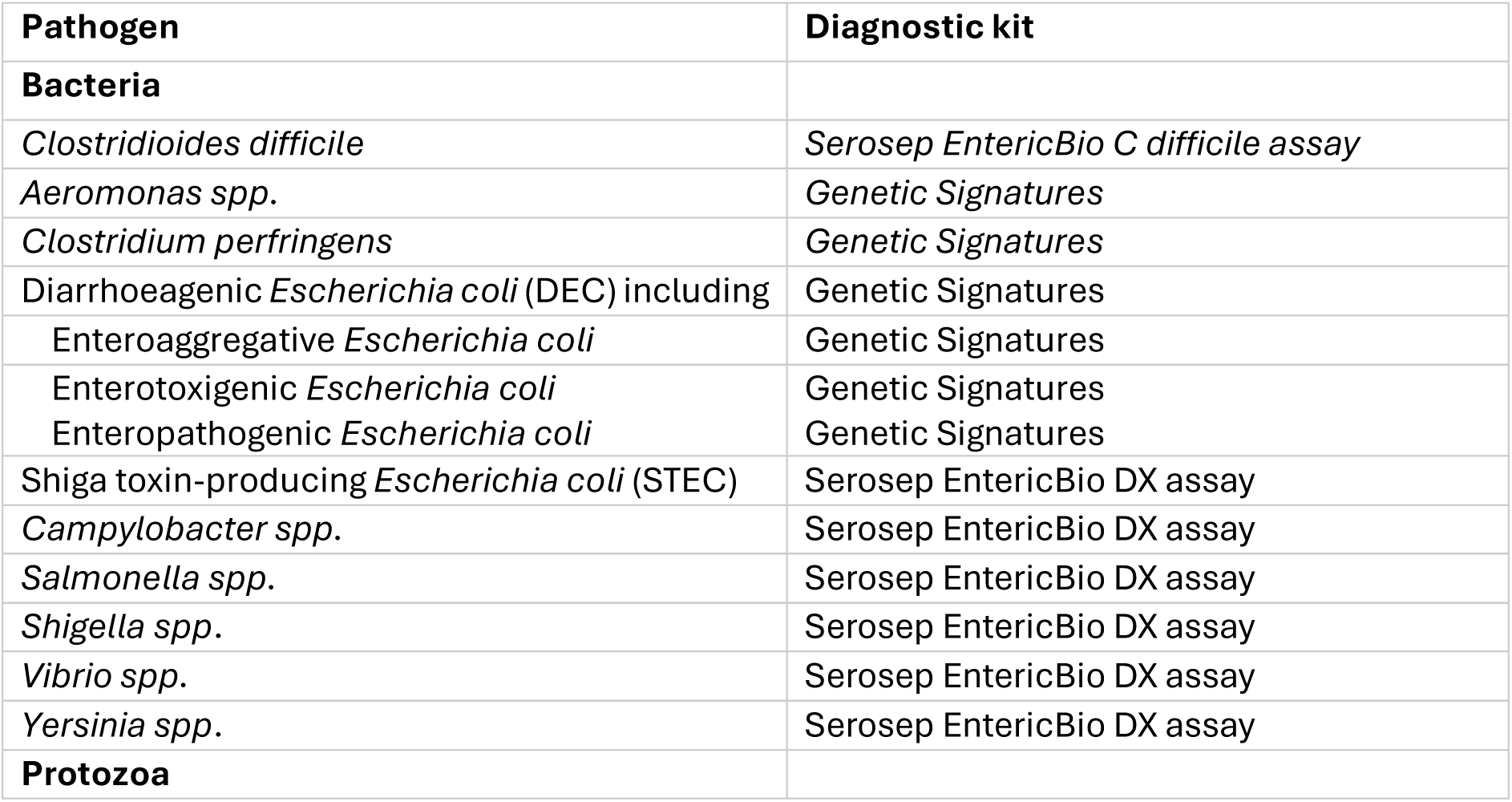
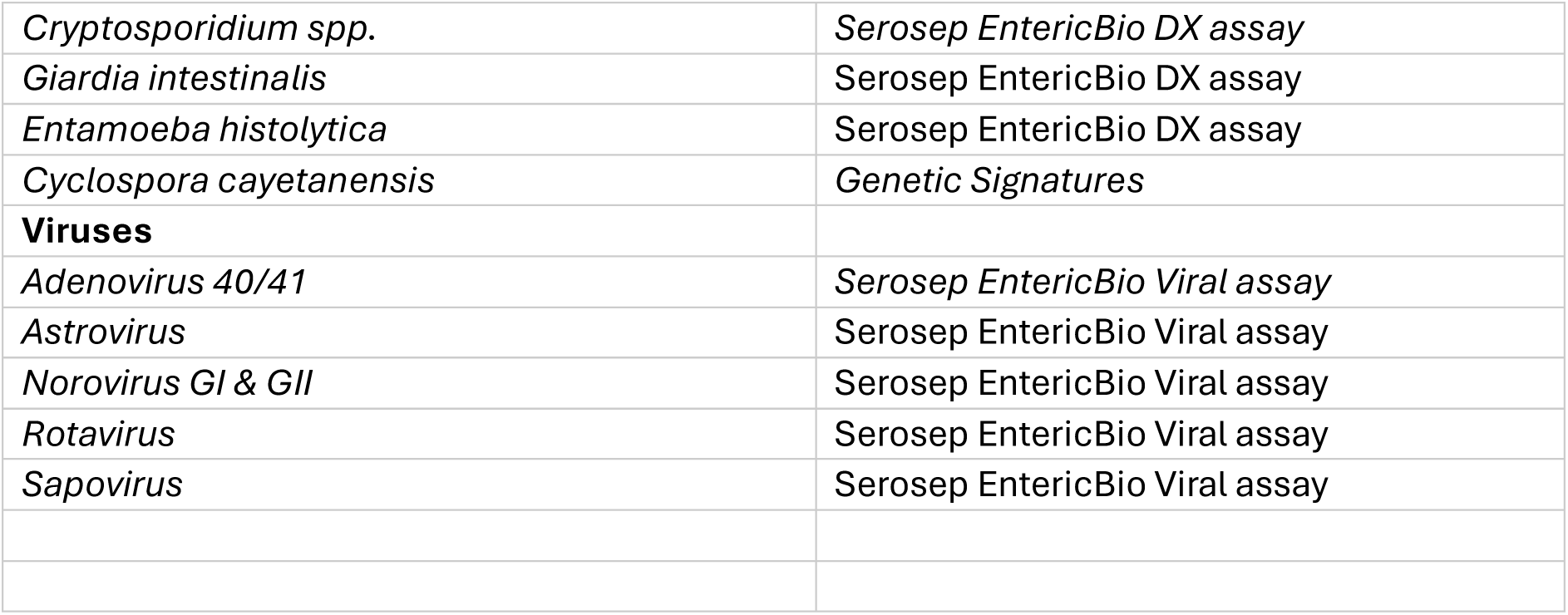
Assays used to detect 21 causative organisms stool specimens.

All specimens are additionally tested for an extended panel of organisms using PCR tests based on a commercial Genetic Signatures 3 base ® Technology, which was evaluated in the routine diagnostic setting at LCL, using commercially-provided pathogen controls, reference isolates and clinical samples, targeting *Clostridium perfringens, Cyclospora cayetanensis* and enabling the inclusion of *Aeromonas spp*., and diarrhoeagenic *Escherichia coli* (enterotoxigenic, enteropathogenic & enteroaggregative strains) into the diagnostic panel.

The GS1 Microlab Nimbus liquid handler system (Hamilton, France) is used with the GS EasyScreen Sample Processing kit for microbial lysis, cytosine conversion and nucleic acid isolation followed by automated PCR setup with GS EasyScreen detection kits. PCR amplification is performed using QuantStudio 5 cyclers (Thermo Scientific, USA). Performance of the EasyScreen tests is verified with bacterial isolates and parasite-positive stool specimens obtained from the reference laboratories, and commercial NATtrol GI Control EQA material (ZeptoMetrix, USA).

Bacterial cultures isolated from EntericBio *Salmonella* or *Campylobacter* reactive samples, or stool specimens with any other reactive EntericBio or EasyScreen target are sent to the relevant reference laboratory for further characterisation: UK Health Security Agency (Gastrointestinal Bacteria Reference Unit, Enteric Virus Unit) or at Public Health Wales (Cryptosporidium Reference Unit). A flow chart showing the testing process is described in Figure 1.

**Figure 1:**
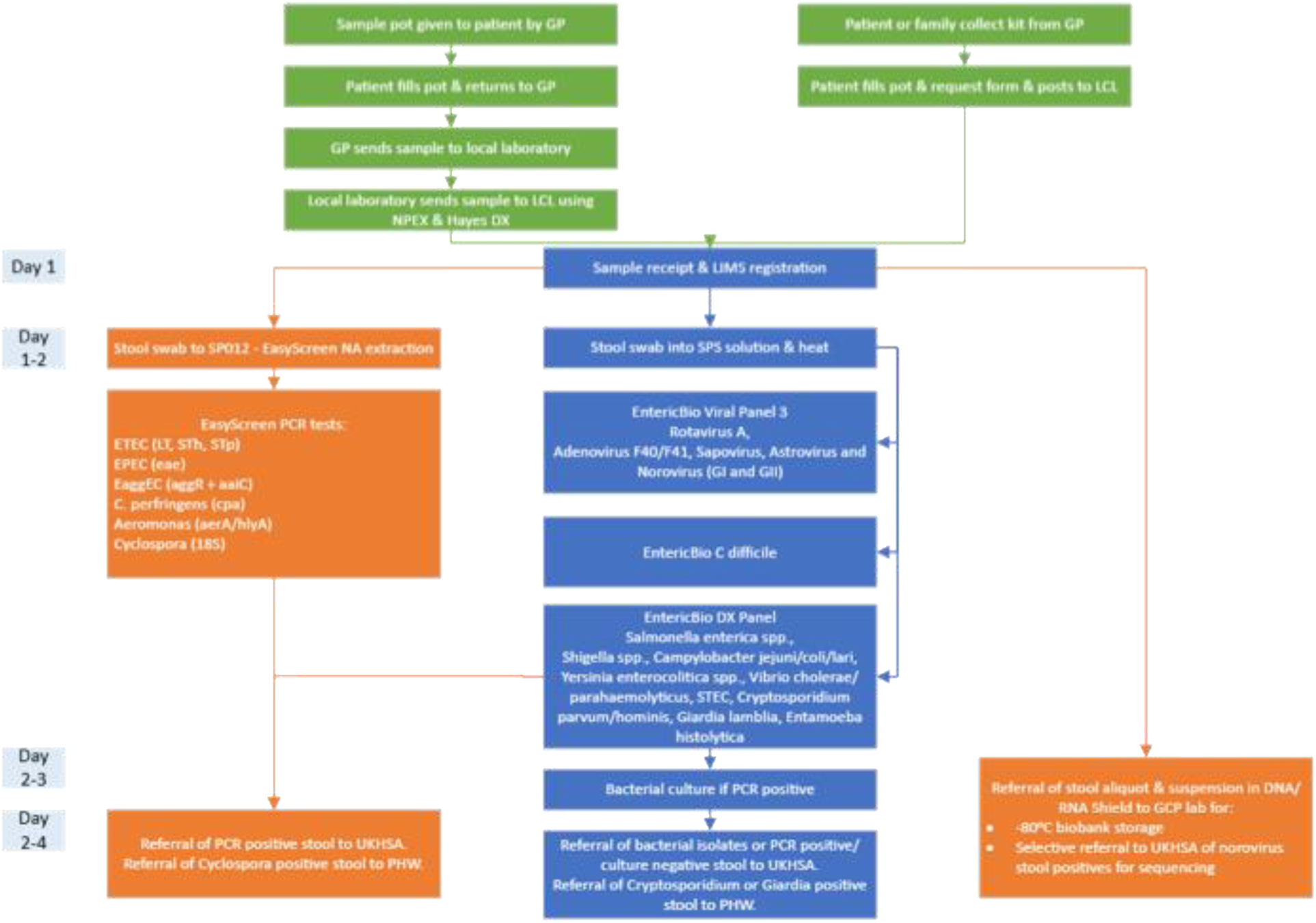
Laboratory testing process for stool samples in IID3 cohorts 1 and 2..

All positive results from cohorts 1 and 2 are reported to the participants’ general practitioners.

### Sample Archiving

Following completion of laboratory testing, any remaining stool specimen for which the participant provided consent for storage, is archived at the University of Liverpool’s Good Clinical Practice Laboratory (GCP) Biobank at -80°C for potential use in future research. Where sufficient material is available, an aliquot is saved in Shield Fluid (Zymo Research, Irvine, USA) to preserve nucleic acid for future molecular applications. Future requests for specimens through the Liverpool Biobank will be overseen by a panel representing the IID3 study partners.

## Bacterial Investigations at GBRU

Bacteria PCR-positive faecal specimens and their bacterial culture isolates are referred to GBRU to undergo one of two workstreams, depicted in Figure 2A.

**Figure 2.**
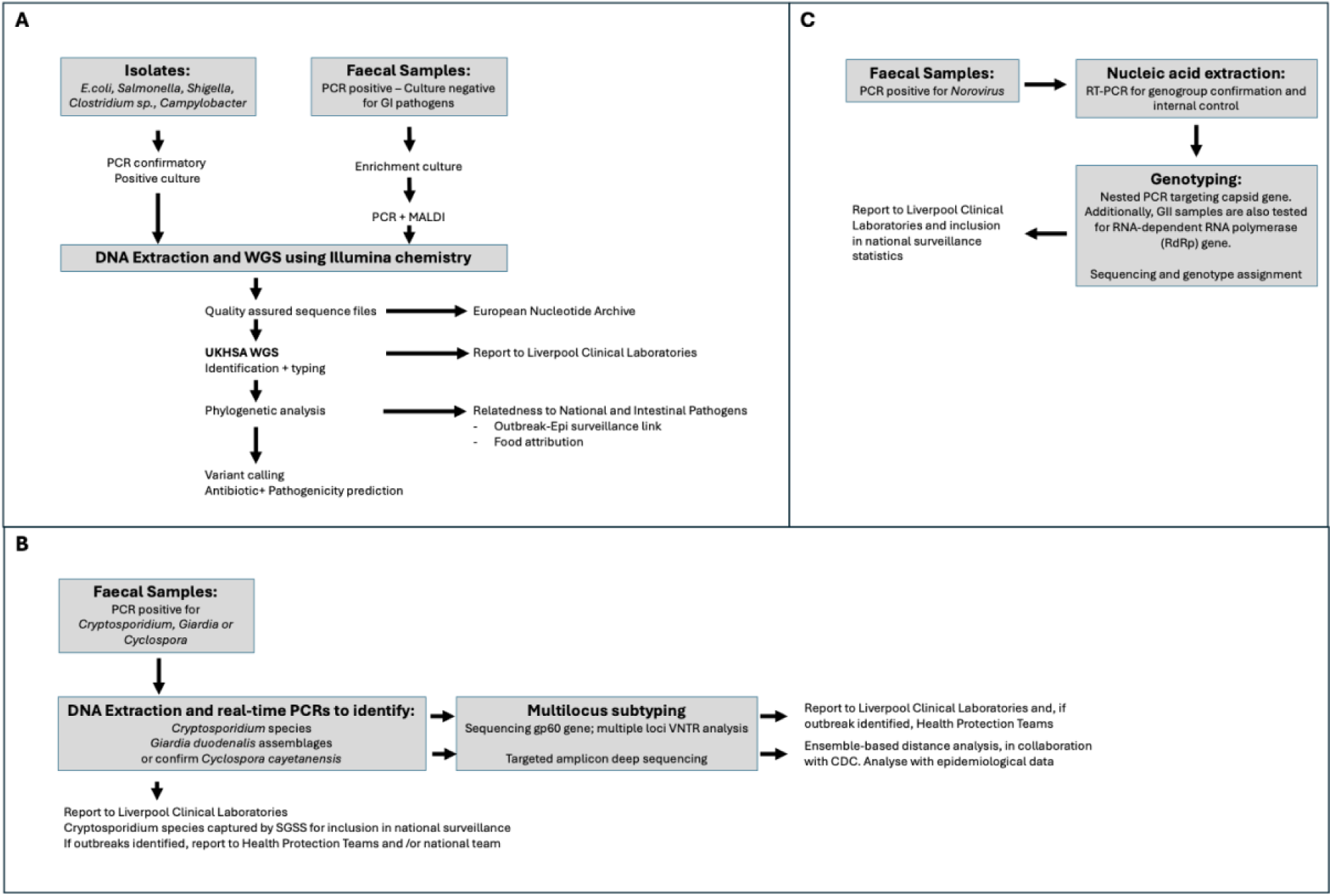
Reference bacteriology workflow of bacterial isolates and faecal specimens referred (A) GBRU, (B) Cryptosporidium Reference Laboratory at PHW; (C) EVU

Faecal specimens are tested by PCR targeting *stx* (encoding Shiga toxin, the definitive characteristic of the STEC group) and *ipaH (*encoding invasion plasmid antigen H, a characteristic of *Shigella* species and the EIEC group), *eae* (intimin gene, a characteristic of the EPEC group), *aggR* (aggregative adherence regulator gene, a characteristic of the EAEC group) and *st* and *lt* (heat-stable and heat-labile toxins, the definitive characteristic of the ETEC group).

Following inoculation of modified tryptone soya broth (mTSB) with a 10 µl loop of faeces and overnight incubation at 37 °C, DNA is extracted from the mTSB enrichment culture using Instagene (Bio-Rad catalogue no. 732–6030). Briefly, 50 µl of enrichment culture is added to 950 µl of sterile distilled water, centrifuged at (11200 g for 1 min) and re-suspended in 200 µl of Instagene reagent. The Instagene suspension is incubated at 56 °C for 30 min, boiled for 20 min, re-centrifuged (11200 g for 1 min) and the supernatant removed to a clean tube for PCR tests.

DNA from faecal extracts is tested using real-time PCR primers and probes targeting *ipaH*, *eae*, *aggR*, *st* and *lt* recommended by the European Union Reference Laboratory (EU-RL) for *E. coli*, at the Istituto Superiore di Sanità, in Rome, on a Rotorgene Q (Qiagen, UK). The amplification parameters are 95 °C for 5 min, followed by 95 °C for 15 s and 60 °C for 60 s. The cycle threshold is set at 0.05 for all targets. Faecal inhibition is monitored by adding the green fluorescent protein (*gfp*) gene as an internal amplification control to the DNA extract. Faecal specimens positive for at least one DEC pathogen target gene (*ipaH*, *eae*, *aggR*, *st* or *lt*) are streaked out in MacConkey agar. Ten morphologically distinct colonies are inoculated into 500 ul of distilled water, boiled for 15 mins and retested using PCR for the appropriate DEC target genes. Individual colonies of *E. coli* testing positive for any one of the virulence genes associated with any of the five DEC groups are genome sequenced to determine sequence type, serotype, virulence profile and genome derived antimicrobial resistance determinants.

Genomic DNA is extracted, fragmented and tagged for multiplexing with Nextera XT DNA Sample Preparation Kits, followed by paired-end sequencing on an Illumina HiSeq platform to produce 80–100 bp short-read sequence fragments (Illumina, Cambridge, UK). Clonal complex (CC), sequence type (ST) and serotype are determined from the genome data, as previously described. MLST assignment is performed using the MOST software (https://github.com/phe-bioinformatics/MOST) (Inouye, et al (2012); Inouye et al (2014)). *In silico* serotyping is performed by mapping FASTQ reads to the genes in the SerotypeFinder – a reference database containing the gene sequences encoding the 180 O antigen groups and the 53 h antigens using Bowtie 2. Only in silico predictions of serotype that matched to a gene determinant at >80% nucleotide identity over >80% length are accepted.

### *In silico* antimicrobial resistance profiling (AMR) profiling

Genome-derived AMR profiles are sought using genefinder (https://github.com/phe-bioinformatics/gene_finder), a customized algorithm that uses Bowtie 2 to map reads to a set of reference sequences and Samtools to generate an mpileup file, as previously described. The presence of resistance genes is defined based on 100% read coverage and >90% nucleotide identity relative to the reference sequence, except for β-lactamase variants, which are determined with 100% read coverage and 100% identity using the reference sequences downloaded from Lahey (www.lahey.org) and NCBI β-lactamase data resources (https://www.ncbi.nlm.nih.gov/pathogens/beta-lactamase-data-resources). Known acquired resistance genes and resistance-conferring mutations relevant to β-lactams (including carbapenems), fluoroquinolones, aminoglycosides, chloramphenicol, macrolides, sulphonamides, tetracyclines, trimethoprim, rifamycins, fosfomycin and colistin are included in the analysis. Chromosomal mutations focused on variations in the quinolone-resistant determining regions (QRDR) s of *gyrA*, *gyrB*, *parC* and *parE*. Reference sequences for acquired and chromosomal resistance genes are curated from those described in the Comprehensive Antimicrobial Resistance Database 1.1.4 (http://arpcard.mcmaster.ca) and the Resfinder 3.2 datasets (https://cge.cbs.dtu.dk/services/data.php).

## Parasitology investigations at the Cryptosporidium Reference Unit

DNA is extracted from stools that are positive for *Cryptosporidium*, *Giardia* and *Cyclospora* using the QIAamp Fast DNA Stool kit (Qiagen) with a pre-treatment in Inhibitex buffer at 95°C for 10 minutes and stored at -20°C until testing.

For *Cryptosporidium*, species are determined by real-time PCR for *C. parvum* using the *Lib13* gene and *C. hominis* using the *A135* gene or PCR-sequencing of the *ssu rRNA* gene [5] and results are reported to LCL. Subtyping is by sequencing part of the *gp60* gene [6] *Cryptosporidium parvum* is further profiled by multilocus variable number of tandem repeats analysis (MLVA) [7,8]

For *Giardia*, assemblages are determined by real-time PCR of the *tpi* gene [9] and results reported to LCL. Assemblage A is further subtyped, as a batch process when sufficient numbers are obtained, using a newly developed real-time PCR based on the *β giardin* gene. This assay is also used to test assemblage A and B negative DNA samples for other assemblages which are confirmed by Sanger sequencing of the PCR product. There is little evidence for the value of subtyping Assemblage B due to the allelic sequence heterozygosity.

For *Cyclospora*, DNA is sent to Centre for Disease Control (CDC) for targeted amplicon deep sequencing and ensemble-based distance analysis [10] (Figure 2B).

## Enteric Virus Reference Laboratory

The characterisation strategy for Noroviruses at EVU-UKHSA involves a reverse transcription step, followed by nested PCR amplifications. Primers specific for GI and GII capsid regions are used for amplification of a genomic region of about 340bp [11,12], that is subsequently sequenced by Sanger Sequencing methodology. All NV GII samples are additionally tested for the polymerase gene using a similar approach [13] (Figure 2C).

### Study timeline

A pilot phase was conducted between January 2023 and August 2023, to test the referral of samples to LCL and the laboratory workflows. Participant recruitment into the main study commenced 01 September 2023 and concluded 31 August 2025.

### Ethics and dissemination

Favourable ethical opinion was granted on the 10^th^ of August 2022 by the East Midlands - Nottingham 1 Research Ethics Committee (IRAS ID 314268, REC Reference 22/EM/0130).

The results will be disseminated to key stakeholders including the FSA, participating general practices and to the academic community through publication and conference presentations. A real-time breakdown of pathogens found in samples received could be viewed on the study [Dashboard](https://app.powerbi.com/view?r=eyJrIjoiNjZiODEwMmUtNWU4Yi00YzJlLTliNjgtYmE1NzA5YWNmNDhmIiwidCI6ImNjOTVkZTFiLTk3ZjUtNGY5My1iNGJhLWZlNjhiODUyY2Y5MSJ9) throughout the study period.

### Outcome and Impact

this collaborative study involving multiple partners has supported the development and application of state-of-the art molecular methods to identify and characterise a broad range of diarrhoea-causing organisms in community-derived faecal specimens. The resulting data will enable calculation of up-to-date pathogen-specific IID incidence estimates in the UK population. Detailed pathogen characterization using methods including WGS will yield additional information on strain types circulating in the community. Faecal samples stored within the GCPlabs at the University of Liverpool will serve as a valuable resource for future study.

## Data Availability

All data produced in the present work are contained in the manuscript

## Authors’ contributions

The study was conceived and designed by NAC, SOB, AM, DH, RC, CJ, GG, MH.

RC, CJ, CB, KE, AH, MMD, ACD, GG, SB, MH, EC.-O. were responsible for the acquisition, analysis, or interpretation of data.

Drafting the work or reviewing it critically for important intellectual content: CB, CJ, GG, KE, CC, RC, DH, MH, NAC, AM,

Final approval of the version to be published: All

Agreement to be accountable for all aspects of the work: All

## Data Statement

Data collected for the study will be de-identified and stored within a secure database by the University of Oxford. It will only be accessible to data analysts who are part of the study team, through a trusted research environment.

## Acknowledgements

We are grateful to all our colleagues at Liverpool Clinical Laboratories, UKHSA, PHW, PHS, PHA, all participating GP practices and the patients who participated in IID3.

## Conflicts of interests

All authors have completed the ICMJE uniform disclosure form at http://www.icmje.org/disclosure-of-interest/ and declare: no support from any organisation for the submitted work.

Nigel Cunliffe is a National Institute for Health and Care Research (NIHR) Senior Investigator (NIHR203756). Nigel Cunliffe, Christina Bronowski Dan Hungerford, Claire Jenkins, Edward Cunningham-Oakes and Alistair C. Darby were affiliated to the NIHR Health Protection Research

Unit in Gastrointestinal Infections at the University of Liverpool, a partnership with the UK Health Security Agency in collaboration with the University of Warwick. Nigel Cunliffe, Christina Bronowski and Dan Hungerford are based at the University of Liverpool. The views expressed were those of the author(s) and not necessarily those of the NIHR, the Department of Health and Social Care, the UK Health Security Agency or Public Health Wales.

## Abbreviation

IID: Infectious Intestinal Disease
IID1: First Infectious Intestinal Disease Study (1993–1996)
IID2: Second Infectious Intestinal Disease Study (2007–2009)
IID3: Third Infectious Intestinal Disease Study (2022-2026)
LCL: Liverpool Clinical Laboratories
UKHSA: UK Health Security Agency
PHA: Public Health Agency
PHS: Public Health Scotland
PHW: Public Health Wales
PCR: Polymerase Chain Reaction
DEC: Diarrhoeagenic Escherichia coli
CRU: Cryptosporidium Reference Unit
EVU: Enteric Virus Unit
GP: General Practitioner
FSA: Food Standards Agency
GCP: Good Clinical Practice
MALDI: Matrix-Assisted Laser Desorption/Ionization
SMAC: Sorbitol MacConkey Agar
TCBS: Thiosulfate-Citrate-Bile Salts-Sucrose Agar
APW: Alkaline Peptone Water
STEC: Shiga Toxin-producing Escherichia coli
EPEC: Enteropathogenic Escherichia coli
EAEC: Enteroaggregative Escherichia coli
ETEC: Enterotoxigenic Escherichia coli
EIEC: Enteroinvasive Escherichia coli
WGS: Whole Genome Sequencing
XLD: Xylose Lysine Deoxycholate Agar
GS: Genetic Signatures
Gfp: Green Fluorescent Protein
EU-RL: European Union Reference Laboratory
CC: Clonal Complex
ST: Sequence Type
MLST: Multi-Locus Sequence Typing
AMR: Antimicrobial Resistance
QRDR: Quinolone Resistance Determining Region
NCBI: National Centre for Biotechnology Information
mTSB: Modified Tryptone Soya Broth
ssu rRNA: Small Subunit Ribosomal RNA
MLVA: Multi-Locus Variable Number Tandem Repeat Analysis
CDC: Centers for Disease Control and Prevention (USA)
tpi: Triosephosphate isomerase gene
GII: Genogroup II (Norovirus classification)
GI: Genogroup I (Norovirus classification)
LUB: Liverpool University Biobank

## Supplementary Figure

**Figure S1.**
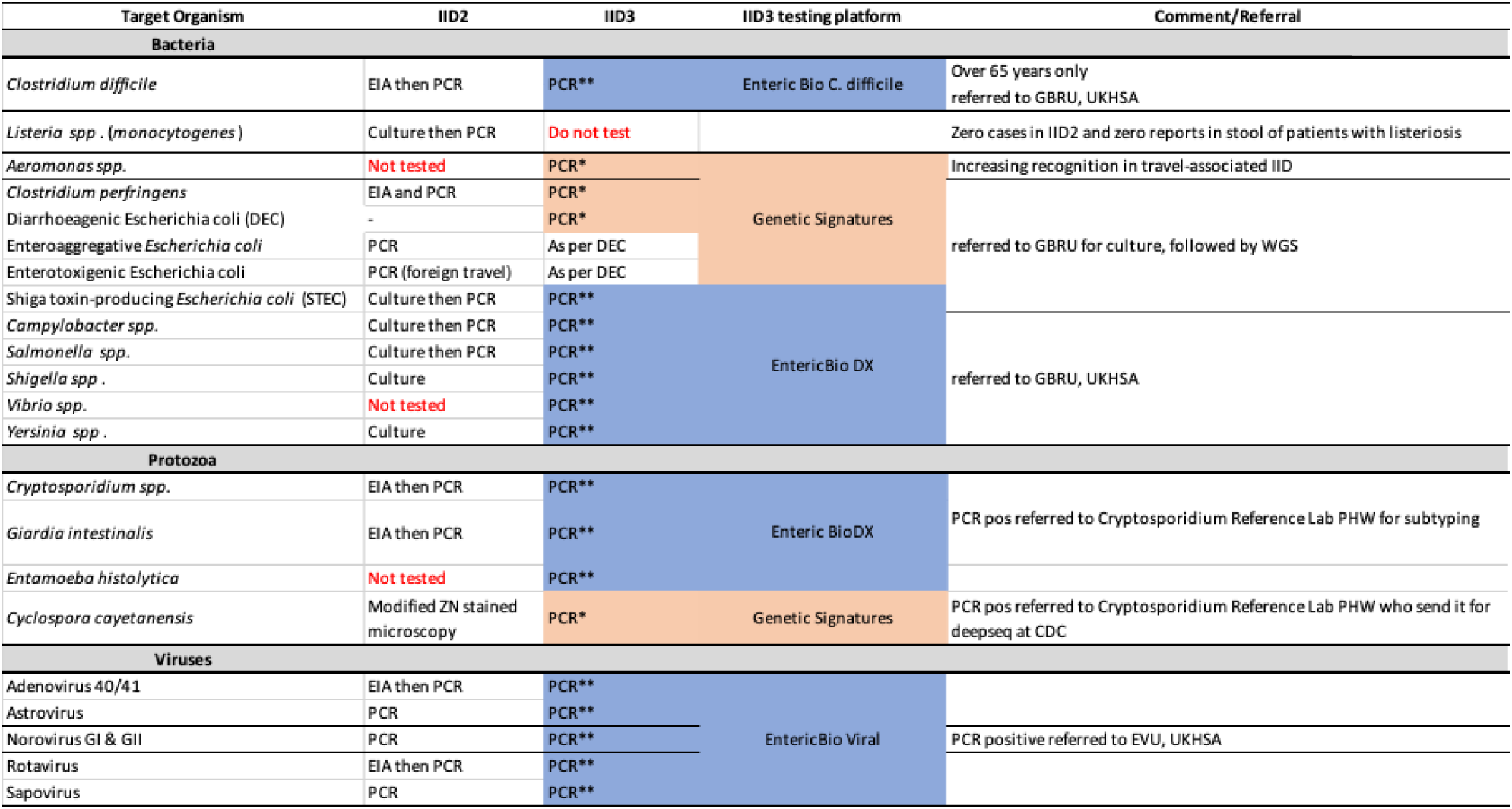
Difference in primary diagnostic testing methods between IID2 and IID3. *bespoke additional target; **included in Serosep Molecular Diagnostics Panels; ***for culture methods see https://www.gov.uk/government/publications/smi-b-30-investigation-of-faecal-specimens-for-enteric-pathogens

